# Consensus based framework for digital mobility monitoring

**DOI:** 10.1101/2020.12.18.20248404

**Authors:** Felix Kluge, Silvia Del Din, Andrea Cereatti, Heiko Gaßner, Clint Hansen, Jorunn L Helbostad, Jochen Klucken, Arne Küderle, Arne Müller, Lynn Rochester, Martin Ullrich, Bjoern M Eskofier, Claudia Mazzà, Mobilise-D consortium

## Abstract

Digital mobility assessment using wearable sensor systems has the potential to capture walking performance in a patient’s natural environment. It enables the monitoring of health status and disease progression and the evaluation of interventions in real-world situations. In contrast to laboratory settings, real-world walking occurs in non-conventional environments and under unconstrained and uncontrolled conditions. Despite the general understanding, there is a lack of agreed definitions about what constitutes real-world walking, impeding the comparison and interpretation of the acquired data across systems and studies. Hence, there is a need for a terminological framework guiding further implementation of digital measures for gait assessment. We used an objective methodology based on an adapted Delphi process to obtain consensus on specific terminology related to real-world walking by asking a diverse panel of clinical, scientific, and industrial stakeholders. Six constituents (‘real-world’, ‘walking’, ‘purposeful’, ‘walking bout’, ‘walking speed’, ‘turning’) have successfully been defined in two feedback rounds. The identification of a consented set of real-world walking definitions has important implications for the development of assessment and analysis protocols, as well as for the reporting and comparison of digital mobility outcomes across studies and systems. The definitions will serve as a common framework for implementing digital and mobile technologies for gait assessment.

## Introduction

Mobility in general, and gait in particular, can be affected by a variety of chronic health conditions, spanning from neurological, such as in multiple sclerosis (MS) and Parkinson’s disease (PD), to respiratory, such as in chronic obstructive pulmonary disease (COPD), cardiac like in congestive heart failure (CHF), or musculoskeletal disorders such as in proximal femoral fracture (PFF) recovery [1–6]. Such functional mobility impairments present a great burden to patients, as they are associated with severe limitations in quality of life [7–9], increased fall risk [10–12], and mortality [13, 14].

Changes in various gait measures such as cadence, gait speed, stride length amongst others may characterize those mobility impairments. The use of digital mobility outcomes (DMOs), which we refer to as digital measures acquired using digital health technology [15] has already been studied in clinical settings using brief standardized tests in a range of diseases [2, 4, 16, 17]. However, a single observation may not be reliable for clinical characterization especially when mobility related disease symptoms fluctuate over time, sometimes even on an hourly basis. Therefore, the objective assessment of gait calls for valid and reliable methods to sensitively capture changes in gait function more frequently [18]. As it is not feasible to increase patient visits to the clinic, a more continuous monitoring outside laboratory or clinical environments is desired [19]. Thus, the continuous assessment of real-world digital measures is essential and opens the opportunity for frequent and long-term remote monitoring [18, 20–22]. In the past years, real-world gait analysis has been technologically enabled by the development of lightweight and easy to use sensor-based systems that can be worn unobtrusively. Although DMOs quantified from real-world data are able to discriminate and detect gait impairments in various diseases [20, 23–26], accepted and routinely used tools are not applied in practice yet [27].

Whilst real-world measurement of mobility holds promise, one fundamental reason for the missing adoption is the difficulty of comparing DMOs across studies due to the inconsistent use of terminology. As an example, a broad variety of terms describing the “real-world” context exist, including “real-life”, “daily-life”, “everyday-life”, and “free-living” [19, 20, 28, 29], that are used interchangeably with ambiguous definitions. This can lead to different test paradigms being considered impeding comparability across measurements, systems, and studies. Furthermore, observed DMO variations may not only be caused by disease symptoms but also environmental factors and measurement protocols, which affect the reliability of DMO assessment. Therefore, unified definitions of relevant DMOs and the context of measurement are necessary to guarantee clinical meaningfulness. As a further example, the term “walking bout” has been used in the context of real-world gait analysis and refers to the quantification of continuous periods of free-living walking [30]. However, walking bout definitions are inconsistent and may include different walking bout durations and number of strides [10, 12, 28, 30–32]. The duration of resting periods between walking bouts [33], and whether turning is considered as part of walking [34, 35] are treated differently as well. However, a clear definition of a “walking bout” is critical, since it directly affects digital measures [28, 36]. Additionally, “turning” needs to be considered as a main constituent of walking, as an average of more than 60 turns per hour has been reported for real-world walking [34]. Due to their high occurence, turnings are likely to break sequences of straight walking into smaller walking bouts. Therefore, the specific definition of a turn directly influences the distribution of walking bouts with regard to their duration. Furthermore, spatio-temporal parameters during straight walking and turning differ [37], such that real-world DMOs based on averages of those parameters strongly depend on whether turning is included in their estimation. Currently, different operational approaches exist for the detection of turning. As an example, turning characteristics may be based on stride to stride angular parameters using foot rotation [38] or on angular changes related to the trunk rotation [34]. This diversity highlights the need for defining “turning” in a framework of DMO assessment, which will anable a more specific operationalization of real-world DMOs.

Overall, a guiding framework for the implementation of DMOs in real-world settings is still missing. Accordingly, the aim of this study was to build a terminological framework in order to drive the development and assessment of DMOs for real-world monitoring. We aimed to reach agreement upon a set of narrative definitions within the scope of the Mobilise-D project [15], which is a five-year EU-funded IMI consortium that will build a technically and clinically-valid system for real-world digital mobility assessment across multiple populations with the goal to improve healthcare.

In this study, we used an objective and systematic consensus process based on an adapted Delphi method [39]. Our results will enable operational definitions to implement mobility assessment algorithms, foster comparability across studies, and serve as a common communication framework for the scientific community. Perspectively, consensus on such a terminological framework is a prerequisite for the adoption of validated digital biomarkers characterizing mobility impairments in various diseases [15, 27].

## Materials and methods

Our approach of defining a terminological framework consisted of the following steps. First, relevant domains and key terms related to real-world walking for the consensus process were identified. Six terms related to four domains of real-world walking needing group consensus were selected (Table 1). For some terms, different aspects were regarded. We proposed a physiological definition of “Walking” and highlighted its relationship to walking bouts. For the definition of “Real-world”, we defined fundamental characteristics, how it is discriminated from standardized measurements and which test paradigms in the clinical context may be regarded as real-world assessment. “Walking speed” has been physically defined. Additionally, we defined the need to consider different granularities when calculating aggregated speed measures and proposed that real-world walking speed needs to be inherently connected to walking bouts. We proposed initial definitions for those eleven aspects based on the study team’s expert knowledge and literature. Iterative feedback was included to improve structure and content of the definitions (for the initial definitions, see S1 Table). We used these definitions as starting point for the subsequent consensus process.

**Table 1.**
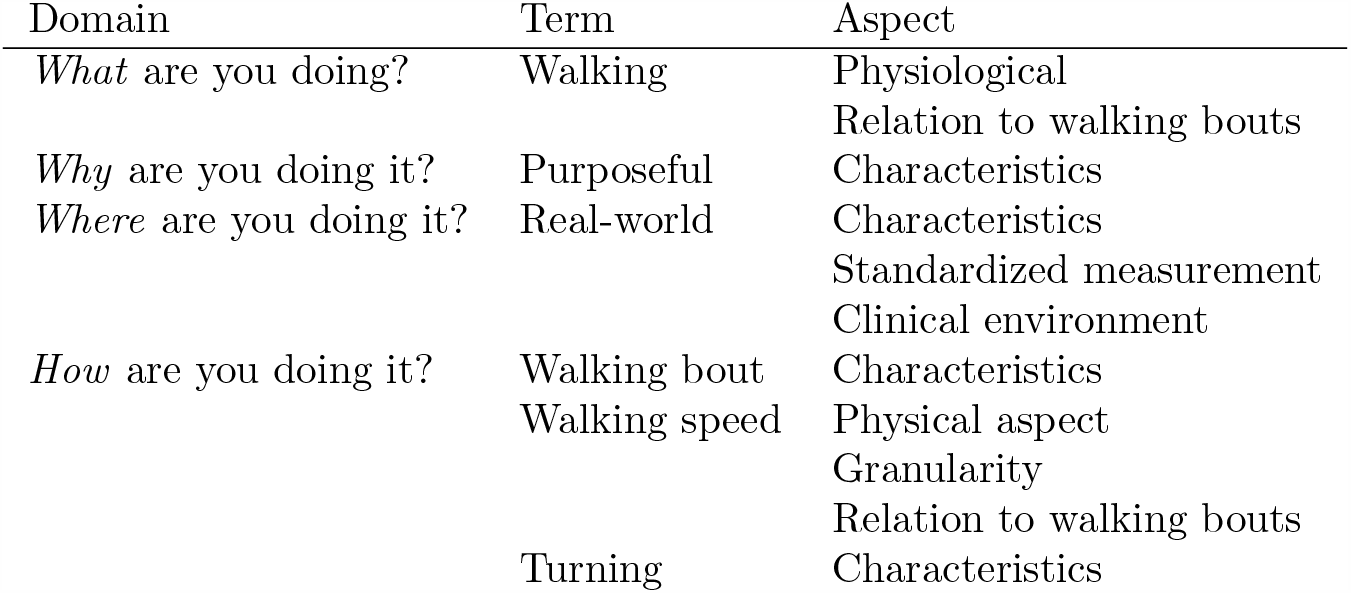
Identified domains and terms needing consensus.

We adopted an objective consensus building process based on the Delphi methodology [39]. In contrast to quantitative methods such as systematic reviews or meta-analyses, which are based on available literature and studies, the Delphi process allows to obtain consensus among experts by determining the level of agreement on a given topic [39]. Specifically, the Delphi method is characterized by anonymity to avoid dominance of single experts, multiple iterations, and feedback to the group. As such, a basic Delphi technique can contain any type of self-administered questionnaire with no meetings [40], which is the approach that was used in this explorative study to quantitatively assess the agreement on the initial definitions.

A consensus process may consist of multiple rounds until agreement on the definitions is reached. Based on a 5-point Likert scale (Fig 1), agreement was quantitatively assessed amongst the participants [41]. A neutral statement (“No opinion”) was included. In our study, agreement to a given definition was defined a-priori as more than 75 % of the answers belonging to the categories “Somewhat agree” or “Strongly agree” [40–42] . In the first round, the participants were asked to independently rate all eleven statements across the six key terms “Walking”, “Purposeful”, Real-world”, Walking bout”, “Walking speed”, and “Turning”.

**Fig 1.**
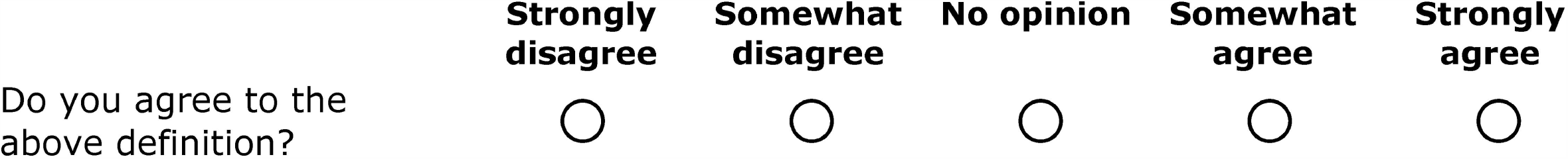
Five-point Likert scale for assessment of agreement.

Additionally, participants were asked to provide free-text comments for each item in order to capture input for the improvement of definitions [41]. In subsequent rounds, we presented modified definitions that previously did not reach agreement and assessed the agreement.

The consensus process was performed among member of the Mobilise-D consortium. The project includes technical, clinical, and industrial expertise of 34 partners from Europe and the USA. All 162 members of the consortium were asked for participation via email. Participants who did not respond in the first round were not invited to participate in the second round. We did not define any exclusion criteria. Data on participants’ “technical or clinical background”, “gait expertise”, “free-living expertise”, and “expertise with patients” were collected in the first round to analyse the panel’s background.

The consensus process was implemented as a series of questionnaires based on the “Survey” feature of the ILIAS e-Learning platform (version 5.4.5, ILIAS open source e-Learning e.V.). It allowed the anonymous acquisition of responses. The participants’ email addresses were linked to access codes, which were provided to start the questionnaire. The use of access codes allowed sending reminders to the participants and preventing double participation. The acquisition and analysis of the data was anonymous. Descriptive statistics was used to investigate participants’ background information and agreement responses in each round. Analyses were conducted using R version 4.0.3 [43]. The code is available on https://doi.org/10.5281/zenodo.4316739. The datasets generated and analysed in the study are available on https://doi.org/10.5281/zenodo.4316564.

Ethical approval for this study was granted by the ethics committee of the University Hospital Erlangen (Re.-No. 241 19 Bc). All participants provided informed consent for inclusion in the study. Participation in this study was voluntary. All data were handled in accordance with European data protection regulations.

## Results

### Consensus process

In total, the consensus process required two rounds until agreement on all definitions was reached. 162 members of the Mobilise-D consortium were asked to participate in the first round of the consensus process. Of those, 79 individuals started the questionnaire. Eight individuals did not sign the participation or data usage agreement and five participants did not complete the questionnaire. Hence, their data was discarded. Data from the remaining 66 participants (40.7 % response rate in the first round) were analysed. Of the participants who completed the first round, 55 individuals started the second round. One individual did not sign the participation agreement and three individuals did not complete the questionnaire. We analysed the data of the remaining 51 participants (continuation response rate of 77.3 %). The overall response rate regarding both rounds was 31.5 %. The professional background of the panel was diverse but homogenously distributed across clinical and technical disciplines (Table 2). Only 12.1 % stated to have no experience in gait analysis. Two thirds of all individuals had expertise in free-living mobility. Most participants (88.7 %) stated to have expertise with patients. As answering the background questions was not obligatory, the total number differs from the total number of individuals who participated in round one of the process.

**Table 2.**
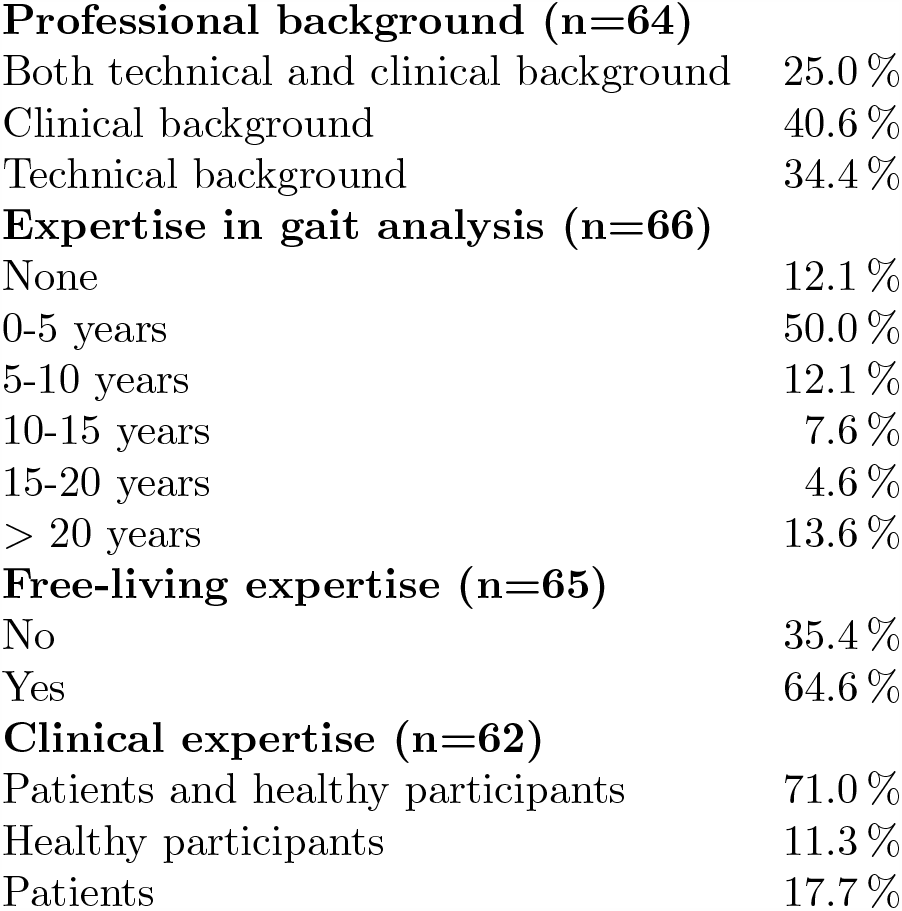
Participant background assessed in the first round.

### Agreement on definitions

In round one, the definitions of “Purposeful” and “Walking speed (Relation to walking bouts)” did not reach agreement (Table 3) and were subject to modification based on participants’ feedback. Although the definition of “Walking bout” reached agreement, there was inclarity regarding its inherent connection to the “Walking speed” definition. “Walking speed” initially assumed a different number of strides required to assess average walking speed (for the initial definitions, , see S1 Table). Therefore, we decided to harmonize the “Walking bout” and “Walking speed (Relation to walking bouts)” definitions, which were both put to vote again in the second round. Full consensus for all definitions was reached in round two (Table 3) resulting in a final set of definitions for real-world gait analysis (Table 4).

**Table 3.** Proportion of agreement and disagreement [%] of definitions. In round two, only those definitions were evaluated, which did not reach agreement in the first round. The lower limit of agreement was a priori defined as 75 %.

| Term | Aspect | Disagreement | No opinion | Agreement | Consensus |
| --- | --- | --- | --- | --- | --- |
| **Round one** |  |  |  |  |  |
| Walking | Physiological | 6.1 | 3.0 | 90.9 | yes |
|  | Relation to walking bouts | 0.0 | 3.0 | 97.0 | yes |
| Purposeful | Characteristics | 10.6 | 21.2 | 68.2 | no |
| Real-world | Characteristics | 4.6 | 1.5 | 93.9 | yes |
|  | Clinical environment | 4.6 | 1.5 | 93.9 | yes |
|  | Standardized measurement | 4.6 | 3.0 | 92.4 | yes |
| Walking bout | Characteristics | 15.1 | 6.1 | 78.8 | yes |
| Walking speed | Physical definition | 1.5 | 1.5 | 97.0 | yes |
|  | Granularity | 1.5 | 6.1 | 92.4 | yes |
|  | Relation to walking bouts | 15.2 | 24.2 | 60.6 | no |
| Turning | Characteristics | 3.0 | 3.0 | 94.0 | yes |
| **Round two** |  |  |  |  |  |
| Purposeful | Characteristics | 15.7 | 5.9 | 78.4 | yes |
| Walking bout | Characteristics | 11.8 | 5.9 | 82.3 | yes |
| Walking speed | Relation to walking bouts | 15.7 | 7.8 | 76.5 | yes |

**Table 4.**
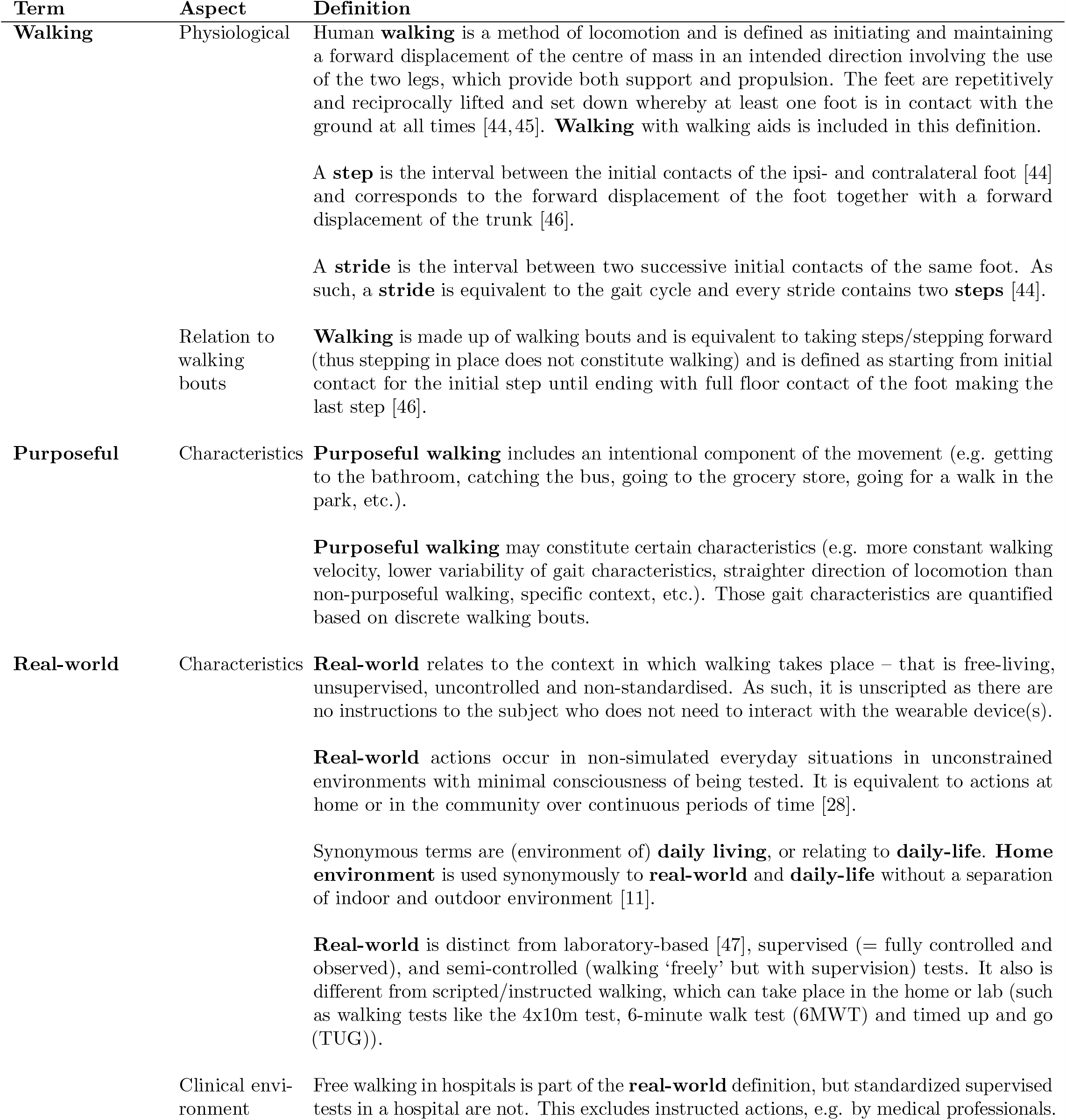
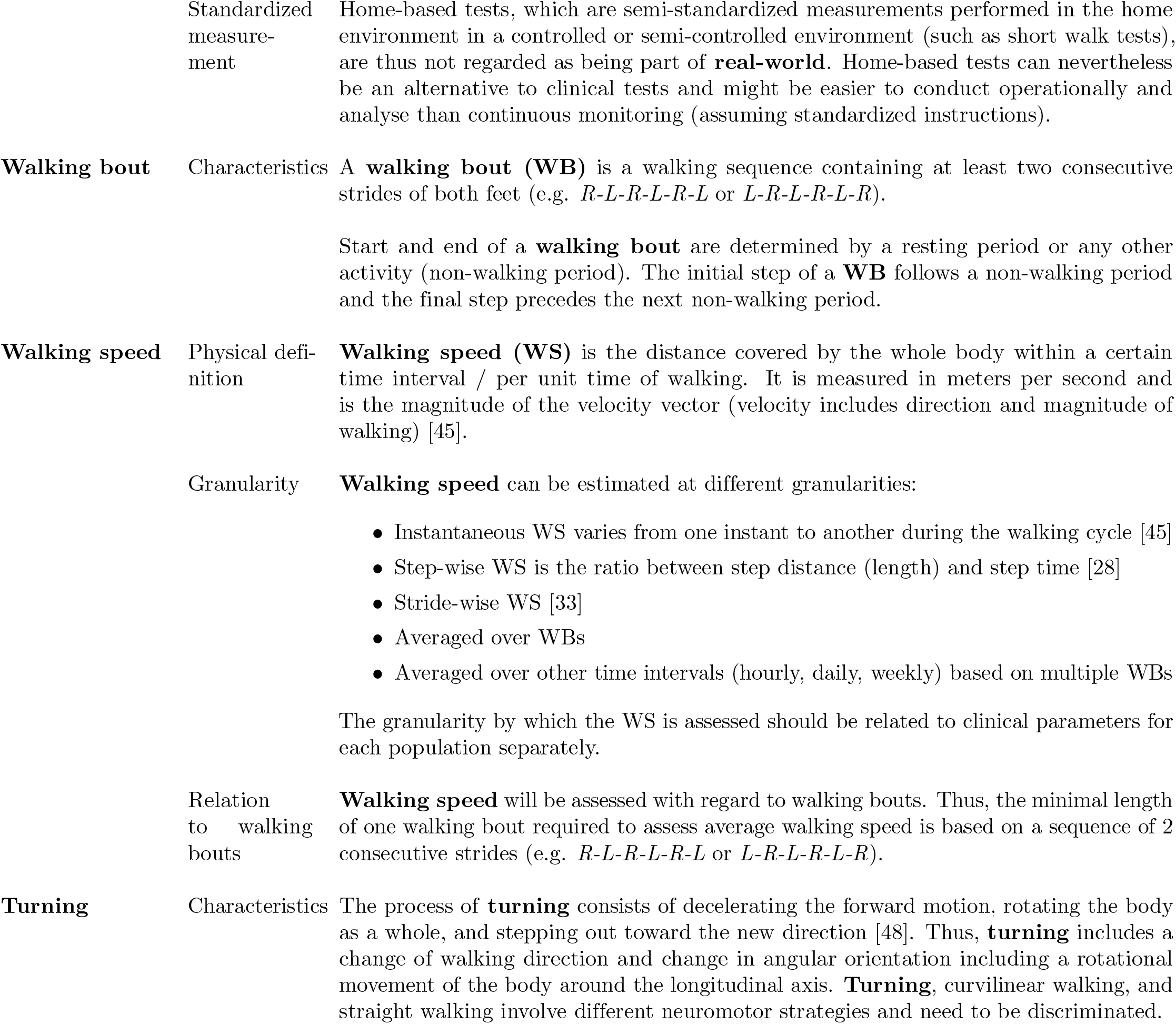
Agreed definitions of terms related to real-world walking. Term Aspect Definition

## Discussion

To the best of the authors’ knowledge, this has been the first study to engage clinicians, academic researchers, and industry stakeholders working in the field of digital gait and mobility measures development to identify and agree upon a framework of narrative definitions for the assessment of DMOs acquired in real-world conditions. An adapted Delphi consensus process allowed to achieve consensus on eleven statements related to six key terms of real-world walking.

A broad definition of “Walking” may include various displacements of the body in space (e.g. for-, back-, or side-ward walking). We, however, define walking to be only associated with forward displacement using both legs in order to assure reliability of DMO assessment in various contexts. Stepping on the spot, side stepping, and backward walking have thus been excluded from this definition. Walking is also not defined in terms of a specific speed. The use of walking aids has been included into the definition as they may be an essential requirement for safe locomotion of people with gait impairments. Otherwise, certain patients and elderly individuals might be excluded from the DMO assessment. We acknowledged steps and strides as basic elements of walking as previously suggested [49]. Furthermore, the definitions include that walking is always made up of walking bouts. It was agreed that walking bouts represent sequences containing at least two full consecutive strides of both feet without a break (e.g. *R-L-R-L-R-L* or *L-R-L-R-L-R*, with *R/L* being the contacts of the right/left foot with the ground, respectively). Start and end of a walking bout are determined by a break that can either consist of a resting period, turning, or any other non-walking real-world activity. More specifically, the start is always defined by an initial step of a walking bout following a non-walking period, while the final step precedes the next non-walking period. Walking bouts are thus an important building block in the terminology framework for the assessment of DMOs acquired in real-world conditions.

“Walking speed” has been referred to as sixth vital sign, as slow walking has been associated with morbidity, cognitive decline, and fall risk amongst others [13, 50]. Despite this, there is still no accepted common measure of mobility that serves across multiple conditions, which is underlined by a wide range of inconsistent testing procedures. With an operative definition of walking speed and a proposition of respective aggregation levels at which it is measured, we aim to provide a common framework to be used across clinical conditions. The physical definition of the walking speed definition reached high consensus. Furthermore, the panel agreed that walking speed will be assessed based on a minimal number of consecutive strides. According to clinical questions, walking speed needs to be assessed with regard to different aggregation levels (hourly, daily, weekly, etc.). This definition is in line with the walking bout definition. On the one hand, this specification yields a unified approach of assessing speed in our framework, but requires a stride-wise analysis of walking, which might not be feasible in all analysis cases, for example when the extraction of single strides is not feasible. The definition also suggests that walking speed derived from strides that are not part of walking bouts (i.e. short strides, shuffling, turning, etc.) should not be considered for the estimation of real-world walking speed.

Daily mobility does not only contain straight walking but also curved walking and turns. Therefore, we included a definition of “Turning” in the framework to guide the implementation of walking bouts and the related DMO assessment. Turning can be regarded as being a deceleration of the forward motion, rotating the body as a whole, and stepping out toward the new direction [48]. It results in a change of walking direction and change in angular orientation including a rotational movement of the body around the longitudinal axis. As an example, a threshold on the rotation angle (e.g. *>* 45^*°*^) at a certain turn duration (e.g. between 0.5 s and 10 s) can be used to detect a turn [34]. This definition does not include all the required aspects for quantitative ambulatory mobility measurement. For example, further discussion will be necessary to define specific angular thresholds between straight and non-straight (i.e. curvilinear) walking. However, those operational definitions are not part of the narrative framework considered in this study and are, therefore, part of future evaluations based on real-world data from different clinical populations.

The environmental context of walking greatly influences DMOs. Thus, it was deemed necessary to specify inclusion and exclusion criteria of what is considered “Real-world”. The participants agreed on different aspects of the terminology: real-world is conceptualized as free-living, unsupervised, uncontrolled, and non-standardized. In the real-world context, the measurement of DMOs should not interfere with daily activities of the participant. The measurement process of real-world DMOs should thus be as non-obtrusive as possible. Accordingly, this definition is distinct from laboratory-based [47], supervised (fully controlled and observed), and semi-controlled (walking freely but with supervision) environments, in which observer and instruction effects might occur and influence DMOs. For example, walking happens in non-simulated real-world situations in unconstrained environments equivalent to actions at home or in the community over continuous periods of time [28]. Daily-living, including home and clinical environment are equivalent to real-world as long as the walking happens unsupervised. Scripted walking capacity tests such as 4×10 m walking conducted at home are excluded from the definition, as significant differences between DMOs derived from those tests and DMOs acquired during unscripted real-world walking are expected [51]. However, differences and relationships between standardized tests and real-world assessments still need to be evaluated in future research studies.

The participants agreed to the definition of “Purposeful” as a consistent term for the assessment of DMOs acquired in real-world conditions. Purposeful walking includes an intentional component of the movement. We assume that unsupervised walking is per se purposeful and that the intentional aspect occurs especially for long walking bouts and needs to be evaluated taking for example contextual aspects of walking into account. Differences between purposeful, self-initiated movements, and movements performed in a supervised (and thus not real-world setting) are discussed in detail in [52], and should be further investigated with the consented real-world walking definitions.

The definitions agreed upon in this study are suggestions in order to capture gait analysis characteristics and properties in the real-world environment. The goal to have working definitions for various clinical populations has, on the one hand, resulted in a rather broad definition of walking (i.e. inclusion of walking aids). On the other hand, clarity on specific parts of the definition (i.e. that walking only includes forward locomotion) will allow to implement very specific digital mobility measures without restricting the application cases.

While the Delphi approach is commonly used to obtain broad consensus among experts by determining the level of agreement on a given topic [39], there is always a certain bias. We used purposive sampling under the assumption that members of the Mobilise-D consortium represented experts in the field of real-world gait research. In general, we acknowledge that the choice of participants restricted to the consortium limits generalizability of the results. However, the consortium includes a large group of experts on gait analysis from Europe and the USA. The participants were homogeneously distributed regarding technical and clinical background. Their views were gathered from a wide range of clinical and academic disciplines to equally represent a breadth of expertise. Some participants stated no experience with gait analysis before. However, most of the participants explicitly mentioned having worked in the field of free-living gait analysis. Moreover, the larger proportion of the participants already had experience in clinical gait analysis. Nevertheless, further work is needed to validate the results of our study in light of an even broader international group of gait experts. For example, the survey could be opened to a wider panel to validate and refine the findings. Furthermore, the framework needs to be evaluated with regard to clinical interpretability of the acquired DMOs based on actual real-world data. Overall, given geographical spread, the Delphi consensus method conducted online was an appropriate tool for gathering the different viewpoints as compared to physical discussion rounds.

The low response rates observed in this study, especially in the first round, are typical for consensus processes and have previously been reported [41]. Especially with large sample sizes, low response rates are considered to be a drawback [53]. Specifically to this study, we invited all members of the Mobilise-D consortium to take part in the study, if they could comment on the topic. Some of the participants invited might not have had a real-world gait analysis background or interest and did therefore not participate in the process. As discussed, the analysis of the participants’ professional background showed that the included participants had relevant experience in the field of interest. Furthermore, the final sample size of 51 participants was higher than the lower threshold of 12, which has been regarded as minimal number to ensure reliability of results in a consensus process [54].

Only group feedback in the questionnaires were provided, as individual feedback was not possible due to anonymity. However, the participants were sent an email with their individual responses and comments after completion of a round. This allowed them to reflect on their own ratings in the subsequent round.

One limitation of this study is that the definitions are only of narrative nature. While the obtained definitions have been objectively derived, some may need refinement according to the practical needs to directly guide algorithm implementation (e.g. thresholds on differentiating turning from curvilinear or straight walking need to be derived from further consensus or based on real-world data). Whilst extracting and analysing DMOs, more detailed definitions need to be derived from the initial framework to enlarge the scope and ensure applicability across different technologies and solutions for real-world gait assessment.

This work was conducted as part of the Mobilise-D project [15] with the aim to guide the data analysis process regarding real-world walking analysis with a focus on the assessment of real-world walking speed. It has to be noted, that different ways of assessing real-world mobility exist, such as analyzing daily activity patterns (e.g. daily step count, physical activity, energy expenditure amongst others). Related digital measures are of high interest for some diseases and might benefit from similar terminological frameworks.

In conclusion, consensus on narrative definitions for the assessment of gait related DMOs acquired in real-world conditions was obtained through an objective consensus process, which has important consequences for clinical gait and mobility research, and the implementation of digital measures. Future work within the community may evaluate DMOs based on those definitions, which will improve comparability between studies and settings. The results of this study have thus important implications for the development of standardized analyzis protocols, as well as for the reporting, and comparison of DMOs. Overall, the definitions will allow a more precise use of those terms in future studies, enabling a stronger congruence of clinical, technical, and regulatory activities in this field.

## Data Availability

The datasets generated and analysed in the study are available in the Zenodo repository, https://doi.org/10.5281/zenodo.4316564
The code is available on https://doi.org/10.5281/zenodo.4316739

https://doi.org/10.5281/zenodo.4316739

## Supporting information

**S1 Table. Initially proposed definitions of terms related to real-world walking assessed in round one.**

