## Supplemental Table 1 for "Consensus based framework for digital mobility monitoring"

### Supplementary Information

**S1 Table.** Initial definitions of terms related to real-world walking assessed in round one.

| Term | Aspect | Definition |
|---|---|---|
| **Walking** | Physiological | Human **walking** is a method of locomotion and is defined as initiating and maintaining a forward displacement of the centre of mass in an intended direction involving the use of the two legs, which provide both support and propulsion. The feet are repetitively and reciprocally lifted and set down whereby at least one foot is in contact with the ground at all times [1, 2]. **Walking** with walking aids is included in this definition.<br><br>A **step** is the interval between the initial contacts of the ipsi- and contralateral foot [1] and corresponds to the forward displacement of the foot together with a forward displacement of the trunk [3].<br><br>A **stride** is the interval between two successive initial contacts of the same foot. As such, a **stride** is equivalent to the gait cycle and every stride contains two **steps** [1]. |
| | Relation to walking bouts | **Walking** is made up of walking bouts and is equivalent to taking steps/stepping forward (thus stepping in place does not constitute walking) and is defined as starting from initial contact for the initial step until ending with full floor contact of the foot making the last step [3]. |
| **Purposeful** | Characteristics | **Purposeful walking** occurs in discrete walking bouts and is therefore directly associated to the definition of a walking bout. Purposeful walking does not specify any gait characteristics (i.e. certain walking speed). |
| **Real-world** | Characteristics | **Real-world** relates to the context in which walking takes place – that is free-living, unsupervised, uncontrolled and non-standardised. As such, it is unscripted as there are no instructions to the subject who does not need to interact with the wearable device(s).<br><br>**Real-world** actions occur in non-simulated everyday situations in unconstrained environments with minimal consciousness of being tested. It is equivalent to actions at home or in the community over continuous periods of time [4].<br><br>Synonymous terms are (environment of) **daily living**, or relating to **daily-life**. **Home environment** is used synonymously to **real-world** and **daily-life** without a separation of indoor and outdoor environment [5].<br><br>**Real-world** is distinct from laboratory-based [6], supervised (= fully controlled and observed), and semi-controlled (walking 'freely' but with supervision) tests. It also is different from scripted/instructed walking, which can take place in the home or lab (such as walking tests like the 4x10m test, 6-minute walk test (6MWT) and timed up and go (TUG)). |
| | Clinical environment | Free walking in hospitals is part of the **real-world** definition, but standardized supervised tests in a hospital are not. This excludes instructed actions, e.g. by medical professionals. |

| | | |
|---|---|---|
| | Standardized measurement | Home-based tests, which are semi-standardized measurements performed in the home environment in a controlled or semi-controlled environment (such as short walk tests), are thus not regarded as being part of **real-world**. Home-based tests can nevertheless be an alternative to clinical tests and might be easier to conduct operationally and analyse than continuous monitoring (assuming standardized instructions). |
| **Walking bout** | Characteristics | We define a **walking bout (WB)** as the sequence of walking periods that may be separated by breaks (resting periods). Each walking period is defined as an interval of at least three successive strides.<br><br>Note: We set no specific threshold for the maximum resting period between consecutive walking periods as the change of digital mobility outcomes with the resting period is unclear [7] and should be investigated for each clinical population separately. |
| **Walking speed** | Physical definition | **Walking speed (WS)** is the distance covered by the whole body within a certain time interval / per unit time of walking. It is measured in meters per second and is the magnitude of the velocity vector (velocity includes direction and magnitude of walking) [2]. |
| | Granularity | **Walking speed** can be estimated at different granularities:<br>• Instantaneous WS varies from one instant to another during the walking cycle [2]<br>• Step-wise WS is the ratio between step distance (length) and step time [4]<br>• Stride-wise WS [8]<br>• Averaged over WBs<br>• Averaged over other time intervals (hourly, daily, weekly) based on multiple WBs<br><br>The granularity by which the WS is assessed should be related to clinical parameters for each population separately. |
| | Relation to walking bouts | We assume that the minimal length of one walking period required to assess average **walking speed** for all clinical populations is 10 steps (5 strides). |
| **Turning** | Characteristics | The process of **turning** consists of decelerating the forward motion, rotating the body as a whole, and stepping out toward the new direction [9]. Thus, **turning** includes a change of walking direction and change in angular orientation including a rotational movement of the body around the longitudinal axis. **Turning**, curvilinear walking, and straight walking involve different neuromotor strategies and need to be discriminated. |
